# Pathway-based polygenic score analysis identifies the NRF2-KEAP1 and mRNA splicing - minor pathways as enriched in shared genetic variation between chronotype and bipolar disorder

**DOI:** 10.1101/2024.02.16.24302920

**Authors:** Laura Fahey, Lorna M. Lopez

## Abstract

**Author Summary:** This study investigates shared genetic influences that may contribute to the circadian rhythm disruption and sleep issues in neurodevelopmental and neuropsychiatric conditions. Using polygenic score analysis on large-scale genetic studies of autism, attention-deficit/hyperactivity disorder, schizophrenia, and bipolar disorder, we tested their ability to predict chronotype and insomnia status of participants in the UK Biobank. Our findings reveal that polygenic scores for autism, schizophrenia, and bipolar disorder are associated with an evening chronotype, while polygenic scores for attention-deficit/hyperactivity disorder, autism, schizophrenia, and bipolar disorder are associated with insomnia. Pathway analysis identified the enrichment of shared genetic variation between chronotype and bipolar disorder in the NRF2-KEAP1 and mRNA splicing minor pathways. Previous studies have linked the NRF2-KEAP1 pathway to the pathology of bipolar disorder and schizophrenia. NRF2 and splicing components have been previously reported to be rhythmically regulated by circadian clock genes. These results suggest a potential role for the NRF2-KEAP1 and mRNA splicing minor pathways in mediating circadian rhythm disturbances in bipolar disorder, providing insights into the genetic basis of sleep issues in neuropsychiatric conditions.

It has been postulated that circadian dysfunction may contribute to the sleep problems prevalent in neurodevelopmental and neuropsychiatric conditions. Genetic correlations between numerous pairs of neurodevelopmental or neuropsychiatric and sleep phenotypes have been identified. We hypothesize that this overlapping genetic variation is enriched in certain biological pathways.

We used genome-wide polygenic score analysis to confirm previously reported genetic correlations and pathway-based polygenic score analysis to identify enriched pathways. We created polygenic scores using summary statistics from the largest genome-wide association studies (GWAS) of autism (AUT), attention-deficit/hyperactivity disorder (ADHD), schizophrenia (SCZ) and bipolar disorder (BP). We tested the performance of these polygenic scores in predicting chronotype and insomnia status of UK Biobank participants. For the pathway-based polygenic scores, we restricted genetic variation to SNPs that mapped to genes within 451 pathways from the Reactome database.

Genome-wide polygenic scores for AUT, SCZ and BP were found to be associated with an evening chronotype, and polygenic scores for ADHD, AUT, SCZ and BP were found to be associated with insomnia status. Pathway-based polygenic score analysis identified the *NRF2 KEAP1* and *mRNA splicing minor* pathways as being enriched for genetic variation overlapping between chronotype and BP. For the *NRF2 KEAP1* pathway, the signal is enriched in the subset of genes that function with KEAP1 to regulate *NRF2* expression. Examination of eQTL data pointed to BP associated SNPs within these gene-sets being associated with expression changes of many genes to which they map. A number of these eQTL SNPs were reported to be genome-wide significant for SCZ in previous studies.

These results demonstrate that the overlapping genetic variation between chronotype and BP is enriched in genes involved in the *NRF2-KEAP1* and *mRNA splicing minor* pathways. Animal model and human cell line studies have previously linked the NRF2 pathway to the pathology of BP and SCZ. Additionally, NRF2 and splicing components have been reported to be rhythmically regulated by circadian clock genes. Our results suggest that these pathways could be involved in mediating the disrupted circadian rhythm phenotype of BP.

## Introduction

Neurodevelopmental and neuropsychiatric conditions (NDPCs) encompass conditions that affect brain function, cognitive-processes, emotions and behaviour. Neurodevelopmental conditions manifest in childhood, with the most prevalent ones including autism spectrum disorder (AUT) and attention deficit hyperactivity disorder (ADHD) (1). Neuropsychiatric conditions typically have onset in adulthood and include schizophrenia (SCZ) and bipolar disorder (BP). NDPCs are characterised by their symptomatology, with many of them displaying overlapping symptoms. One example of such overlapping symptoms are sleep problems, considered a core symptom in the diagnostic criteria for BP (DSM-5; American Psychiatric Association, 2013), present in up to 80% of individuals with SCZ (3) and AUT (4,5) and observed in up to 50% of those with ADHD (6).

Sleep is regulated by sleep/wake homeostasis, which balances the urge to sleep and the requirement for wakefulness. As wakefulness persists, the homeostatic sleep drive intensifies, promoting sleep. Sleep is also regulated by the circadian clock, a biological timing mechanism that is controlled by 24-hour transcription-translational feedback loops involving several core clock genes. The central circadian pacemaker in the suprachiasmatic nucleus (SCN) of the hypothalamus responds to external cues, such as light signals, and synchronizes circadian rhythms across all cells in the body (7).

Both sleep/wake homeostasis and circadian rhythm have been shown to be disrupted in NDPCs. Disruption of homeostatic sleep drive, as indicated by reduced slow-wave activity, has been reported in SCZ and early-stage psychosis (8,9). Melatonin and cortisol are two hormones involved in circadian regulation. Melatonin is produced at night, while cortisol is primarily produced in the morning (10). Dysregulation of both hormones has been observed in NDPCs. For example, AUT has being associated with lower levels of melatonin at night (10,11) and ADHD has been associated with lower levels of cortisol after wakening (12). Significant reductions in melatonin levels have also been reported in the cerebrospinal fluid of BP patients (13). Conversely, activities that disrupt circadian rhythms, such as shift work and frequent transatlantic flights, have been associated with adverse mental health (14). Hence, a bidirectional relationship appears to exist, where environmental disruptions to circadian rhythm exacerbate NDPC symptoms, while inherent circadian rhythm disturbances potentially contribute to the development of NDPCs or share underlying mechanisms. This supports the hypothesis that overlapping genetic variation contributes to both circadian disruption and the development of NDPCs. Indeed, mouse models with circadian clock gene mutations display symptoms reminiscent of human NDPCs, with different gene or protein domain mutations resulting in varying sets of NDPC symptoms (15).

Genome-wide association studies (GWASs) have identified hundreds of genome-wide single nucleotide polymorphisms (SNPs) associated with different NDPCs and sleep related phenotypes. Chronotype refers to individual’s preference for wakening and preforming activity earlier in the day (morning chronotype) or later (evening chronotype). It is a behavioural manifestation of an individual’s circadian clock and is often used as a proxy phenotype for circadian timing (16). In modern society, having a morning chronotype is considered advantageous due to its alignment with early school and work start times. Consequently, individuals with an evening chronotype may experience reduced sleep on work/school days and compensate for it on free days - this is known as ‘social jetlag’ (17). Evening chronotypes have being associated with numerous adverse psychological outcomes (18). The most recent GWAS of a morning chronotype reported enrichment of chronotype associated loci in the clock genes *PER1, PER2, PER3, CRY1* and *BMAL1*. Furthermore, the top enriched biological pathways relate to the circadian clock (19).

Surprisingly, no core clock genes have been linked with any NDPC through large-scale GWAS. However, genome-wide correlations in GWAS identified SNP effects have been reported between many sleep traits and NDPCs using the linkage disequilibrium score regression tool (Figure 1). This relationship is negative for the advantageous morning chronotype and positive for insomnia and daytime sleepiness, which are both debilitating. Polygenic score (PGS) analysis can also be used to assess genetic correlations between phenotypes. If a PGS created using effect sizes from a GWAS of one phenotype can significantly predict the phenotype of samples in an independent cohort, it indicates shared genetic variation between the two phenotypes in question (20). PGSs for ADHD and depression have been reported to positively correlate with initiating and maintaining sleep and excessive somnolence in children. Additionally, a PGS for anxiety disorder correlated with nightmares in children (21). These genetic correlation findings suggest that the shared genetic background between NDPCs and circadian rhythms likely extends genome-wide, beyond the core clock genes.

**Figure 1.**
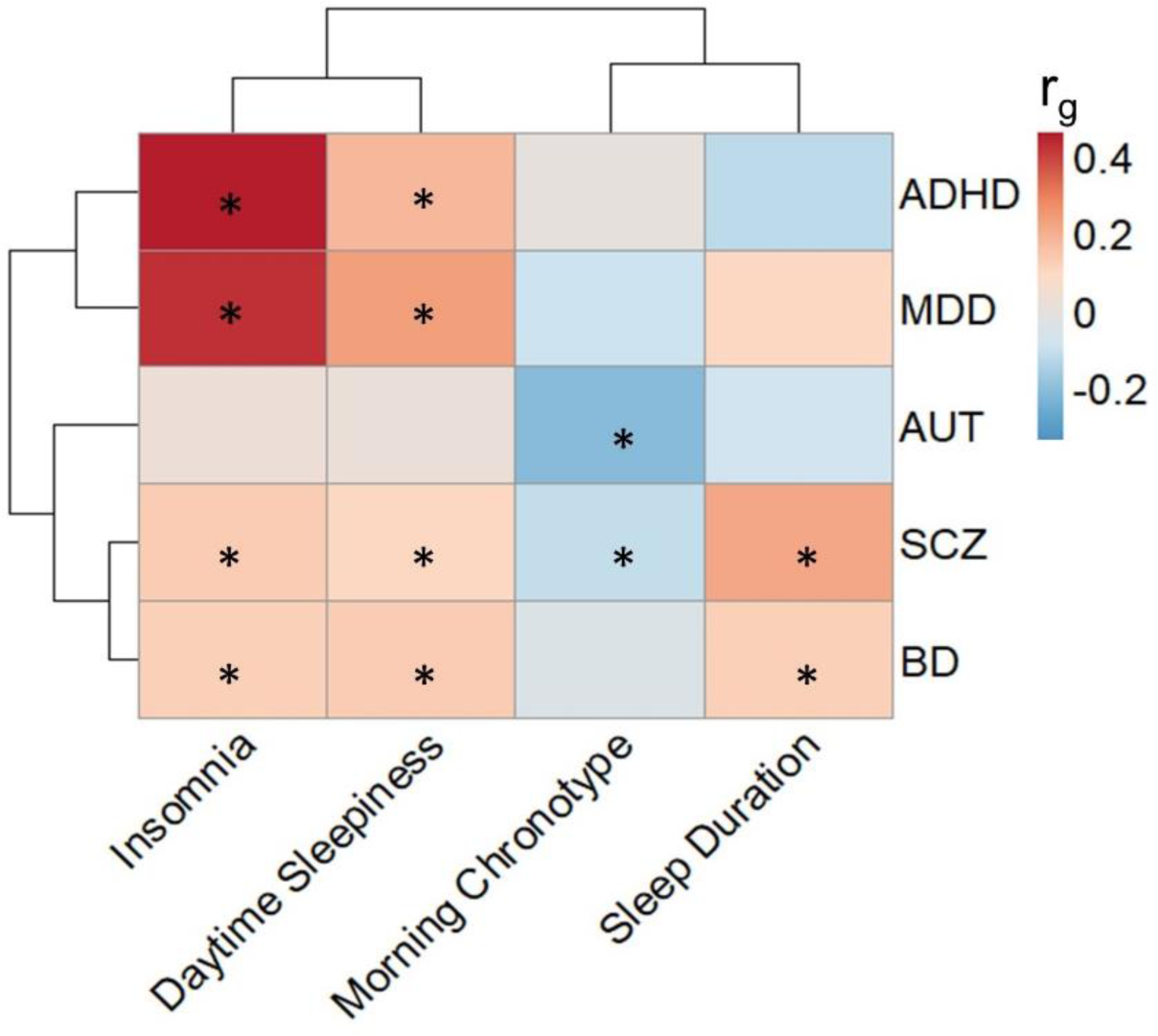
Published genetic correlations (r_g_) between pairs of neuropsychiatric or neurodevelopmental and sleep phenotypes. Heatmap cells marked with an asterisk (*) indicate that they were reported as statistically significant in the original publications, after multiple testing correction. The original publications include the latest and largest genome-wide association studies of Autism (AUT; Grove et al., 2019), morning chronotype (Jones et al., 2019), attention-deficit hyperactivity disorder (ADHD; Demontis et al., 2023), bipolar disorder (BP; Mullins et al., 2021), major depressive disorder (MDD; Howard et al., 2019), sleep duration (Dashti et al., 2019) and daytime sleepiness (Wang et al., 2019). Rows and columns are clustered based on similarity.

Exploring the genes, biological pathways and tissue types impacted by the shared genetic variation between sleep and NDPCs has been a relatively unexplored area. A gene-based cross-trait meta-analysis identified forty-four genes common to both insomnia and ADHD, and found them to be enriched for genes within synaptic-related pathways (22). Another study identified 149 loci shared between psychiatric disorders (SCZ, BP and depression) and sleep traits (chronotype and sleep duration). Forty-nine lead SNPs at these loci were found to act as eQTLs for 115 genes across various brain tissues including the basal ganglia, cortex, hippocampus and cerebellum (23).

The aims of this study were two-fold. First, we investigated if genome-wide polygenic scores for ADHD, AUT, SCZ and BP could predict insomnia and chronotype in UKB. Secondly, using pathway-based polygenic score analysis, we investigated the biological pathways implicated by shared genetic variation between these NDPC and sleep phenotypes. We report that polygenic scores for AUT, SCZ and BP were associated with chronotype status and polygenic scores for all four NDPC phenotypes were associated with insomnia. BP polygenic scores, restricted to genetic variation in the *KEAP1-NRF2* and *mRNA splicing-minor* biological pathways were both significant in predicting chronotype, over and above the background signal. To gain further biological insights, we reviewed expression quantitative trait loci data, and found many BP associated SNPs within these gene sets to be associated with expression changes of the genes to which they map.

## Results

### Genome-wide polygenic score analysis

To test for indication of shared genetic variation, we investigated if genome-wide polygenic scores for AUT, ADHD, SCZ and BP could predict chronotype and/or insomnia status of UKB participants. We implemented this using the PGS tool, SBayesRC, which re-estimates GWAS SNP effect sizes using a model that incorporates external linkage disequilibrium (LD) and biological annotation data (24). As discovery data for PGS analysis, we used publicly available GWAS summary statistics for the NDPC phenotypes, which were based on studies containing between 46,350 (AUT) and 225,534 (ADHD) samples. As target data for PGS analysis, we used UKB genotype and questionnaire data related to chronotype and insomnia. After quality control, we had 409,630 samples with chronotype data and 239,918 samples with insomnia data (see methods).

Polygenic scores for AUT, SCZ and BP, were statistically significant for association with chronotype status (AUT: r^2^ = 3.57×10^−4^ [95% CI = 2.51×10^−4^ - 4.82×10^−4^], p = <2.20×10^−16^; SCZ: r^2^ = 5.13×10^−4^ [95% CI = 3.84×10^−4^ - 6.61×10^−4^], p = <2.20×10^−16^; BP: r^2^ = 2.97×10^−5^ [95% CI = 5.71×10^−6^ - 7.25×10^−5^], p = 4.84×10^−4^) (Figure 2, Supplementary Table 1). Polygenic scores for all four GWAS phenotypes were statistically significant for association with insomnia, the strongest association being between the ADHD PGS and insomnia (ADHD: r^2^ = 2.36×10^−3^ [95% CI = 1.99×10^−3^ - 2.76×10^−3^], p = <2.20×10^−16^; AUT: r^2^ = 3.69×10^−5^ [95% CI = 4.29×10^−6^ - 1.02×10^−4^], p = 2.93×10^−3^; BP: r^2^ = 1.10×10^−4^ [95% CI = 4.19×10^−5^-2.09×10^−4^], p = 2.90×10^−7^; SCZ: r^2^ = 3.32×10^−5^ [95% CI = 3.07×10^−6^-9.52×10^−6^], p = 4.79×10^−3^; Figure 2; Supplementary Table 1). To enable direct comparison to the pathway PGS analysis in the next section, we also report genome-wide PGSs calculated using the tool, PRSice-2 (Choi & O’Reilly, 2019), with a P-value threshold of 0.5 (Supplementary Table 1). These results are very similar to those obtained with SBayesRC, with the exception that SBayesRC performed better in the prediction of an evening chronotype based on all four NDPC PGSs, and also performed better in the prediction of insomnia based on the ADHD PGS. This improvement is to be expected given that the calculation of PGSs using LD clumping and p-value thresholding, as in PRSice-2, is not the optimal method for genome-wide PGS analysis (26).

**Figure 2.**
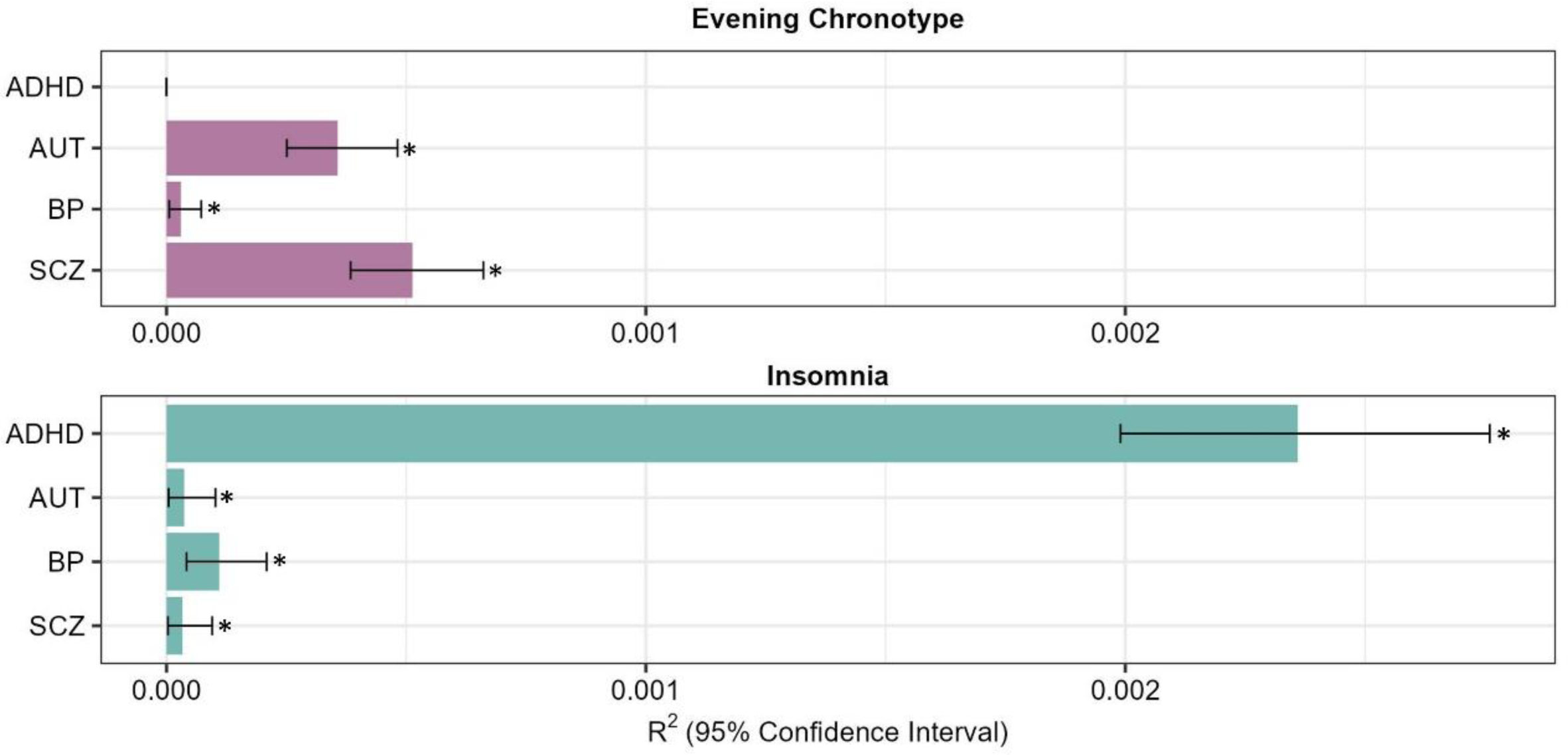
Genome-wide polygenic score analysis results. Linear regressions were run with UK Biobank chronotype or insomnia status as the dependent variable and SCZ, BP, AUT or ADHD polygenic score as the independent variables. UK Biobank phenotypes were corrected for sex, genotyping batch, UK Biobank assessment centre and the top ten ancestry-informative genetic principal components prior to running this regression. Horizontal bars show the variance explained (R^2^), error bars indicate 95% confidence intervals. Bars marked with an asterisk (*) indicate that they surpass the Bonferroni-corrected *p* value threshold of p ≤ 0.0001.

### Pathway-based polygenic score analysis

Pathway-based polygenic score analysis was performed to test if shared genetic variation exists between AUT, ADHD, SCZ or BP and chronotype or insomnia status at the biological pathway level. Here, all eight pairs of phenotypes were tested, regardless of whether statistically significant results were observed at the genome-wide level, because it is possible that shared genetic effects are present in certain biological pathways, but the effects are too small to be observable at a genome-wide level. We tested all Reatome pathways that contained between 50 and 500 genes, resulting in 451 pathways to test. Pathway-based polygenic scores were derived using SNPs that map by position to each of the 451 pathways.

Two pathways were statistically significant for the prediction of chronotype status based on BP genetic variation. These pathways encompass the *mRNA splicing - minor pathway* (r^2^ = 7.5×10^−5^ [95% CI = 3.13×10^−5^-1.26×10^−4^], P = 3.18×10^−8^, competitive P = 1.33×10^−5^) and the *KEAP1 NRF2 pathway* (r^2^ = 7.18×10^−5^ [95% CI = 2.92×10^−5^-1.33×10^−3^], P = 6.26×10^−8^, competitive P = 1.33×10^−5^; Figure 3, Supplementary Table 2). Note that the competitive P value achieved for each of these significant gene-sets was the lowest possible based on the 75,000 permutations performed. Interestingly, the *mRNA splicing - minor* pathway also ranked the highest, in terms of competitive P value, for the prediction of chronotype based on the SCZ PGS, but this did not surpass the multiple testing corrected threshold (r^2^ = 5.62×10^−5^ [95% CI = 1.96×10^−5^-1.12×10^−4^], P = 1.65×10^−6^, competitive P = 1.0×10^−3^; Supplementary Table 2).

**Figure 3.**
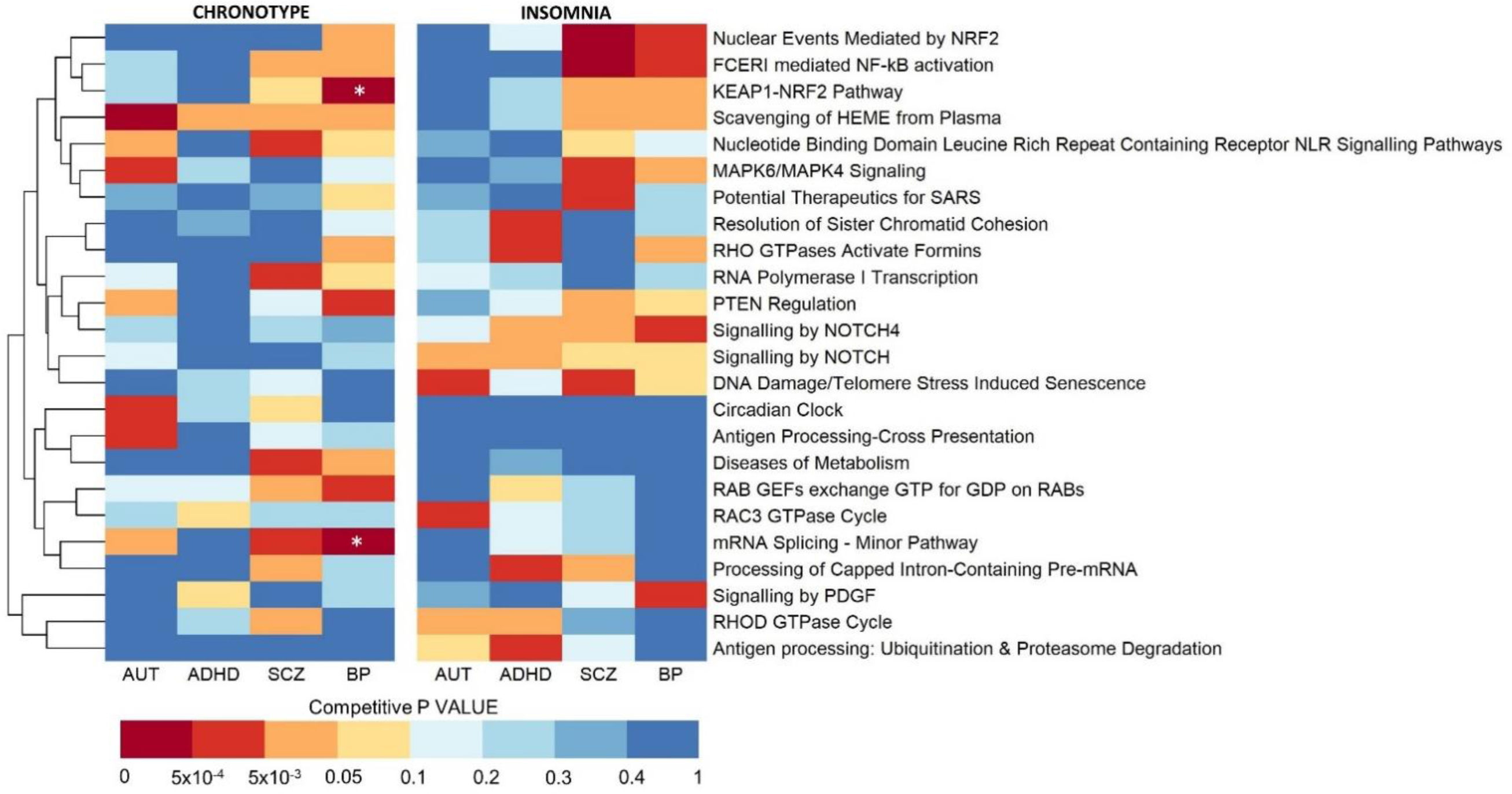
Top performing pathways from pathway-based polygenic score analysis. Performance metrics for pathway-based polygenic score analysis were calculated by regressing insomnia or chronotype case status on each of the pathway-based polygenic scores. Competitive P values were calculated by creating at least 5,000 background polygenic scores, consisting of random sets of post-clump SNPs, the same size as the pathway under test (in this way, pathway size is controlled for). The competitive P value takes into account the number of times a background gene-set obtains a lower null P value than the pathway being tested. The four pathways that resulted in the lowest competitive P values for each of the eight phenotype pairs were identified, and their competitive P values across each of the eight phenotype pairs were plotted as a heatmap. The white stars represent pathway based polygenic scores statistically significant after multiple testing correction (P < 1.39×10^−5^). Rows are clustered based on similarity.

No pathway PGS achieved a competitive P value that passed multiple testing correction for the prediction of insomnia. The pathway *Nuclear Events Mediated by NRF2,* which is a subpathway of *KEAP1 NRF2,* ranked as the second and third highest performing pathways, out of all 451 tested, for the prediction of insomnia based on SCZ and BP PGSs, respectively (Figure 3, Supplementary Table 3). Competitive P values for the top four pathway PGSs for each of the eight phenotype pairs are visualised as a heatmap in Figure 3.

### Post-hoc analyses

#### Interrogations of significant pathway-based polygenic score analysis results

The *mRNA Splicing – Minor Pathway* encompasses 50 genes, to which 518 SNPs map, while the *KEAP1 NRF2* pathway comprises 107 genes, to which 1,356 SNPs map. To explore potential enrichment of the observed signal within specific subsets of these pathways, we investigated Reactome subpathways, some of which were initially omitted due to our criteria of excluding pathways with fewer than 50 genes. Given the relatively small size of the *mRNA splicing – minor pathway*, it does not contain any Reactome subpathways. In contrast, the KEAP1 NRF2 pathway could be broken down into two Reactome subpathways, and an additional pathway that we established comprising of all genes that do not fall into these two subpathways, which we referred to as *Regulation of NRF2 by KEAP1*. Among the *KEAP1 NRF2* sub-pathways, the gene-set that explained the most variance was *Regulation of NRF2 by KEAP1* (R^2^ = 5.0×10^−5^ [95% CI = 1.6×10^−5^-1.03×10^−4^], P = 6.25×10^−6^, competitive P = 1.87×10^−4^; Figure 4a; Supplementary Table 4). To account for the size of the pathways, we plotted the average variance in chronotype explained per SNP for these pathway PGSs, in comparison to the average variance explained per SNP by a PGS constructed using SNPs that map to all HGNC gene symbols (Figure 4b; Supplementary Tables 2 and 4).

**Figure 4.**
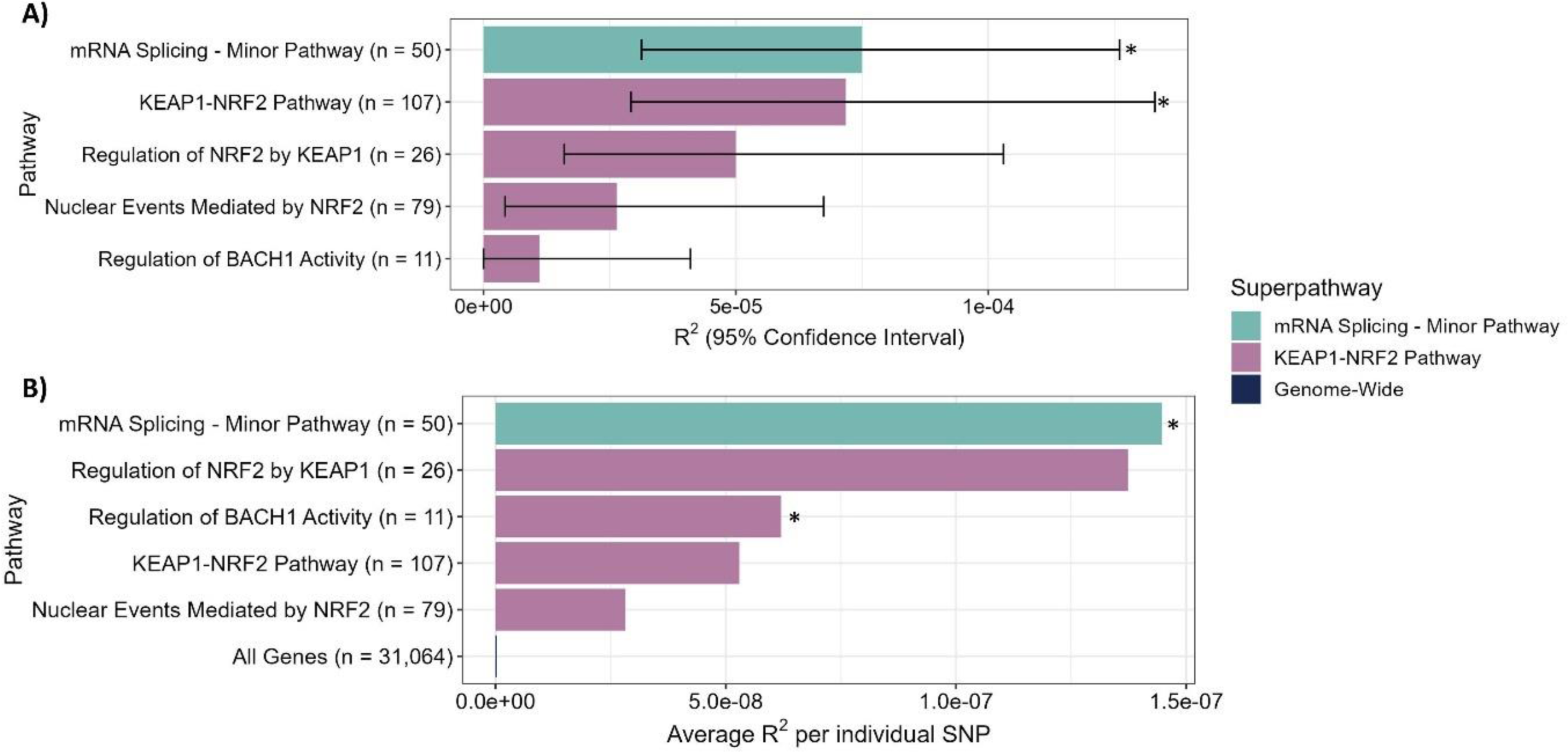
Variance explained (R^2^) by the statistically significant pathway-based BP PGSs, and their subpathways, in predicting chronotype. Bars marked with an asterisk (*) indicate that they surpass the Bonferroni-corrected P value threshold of P ≤ 1.39×10^−5^. A) Total variance explained per pathway. Horizontal bars show the variance explained (R^2^), error bars indicate 95% confidence intervals. B) Average variance explained per SNP. This was obtained by dividing the total variance explained per pathway by the number of SNPs in that pathway.

#### eQTL Analysis Results

To gain a better understanding of the effect of BP genetic variation on the expression of genes within the *mRNA Splicing – Minor Pathway* and *KEAP1 NRF2* pathways, we reviewed expression quantitative trait loci (eQTLs) data in the database, QTLBase (27). For the *KEAP1 NRF2* pathway, five genes satisfied the criteria of having gene P values < 1×10^−2^ for BP and < 1×10^−3^ for chronotype based on gene-based MAGMA analysis. Of these genes, three contained BP SNPs associated with their expression change: *cullin-3* (*CUL3*), *proteasome 26S subunit ubiquitin receptor*, *non-ATPase 2* (*PSMD2*) and *proteasome 20S subunit beta 10* (*PSMB10*).

CUL3 is a core component of the CUL3-dependent E3 ubiquitin ligase complex which mediates ubiquitination and subsequent degradation of target proteins. This complex associates with KEAP1 to ubiquitinate NRF2, maintaining its abundance at a low level (28) At *CUL3*, the BP allele of the most significant BP SNP (rs6748341) is associated with decreased expression of *CUL3* in the brain cortex (P = 4.91×10^−6^). Interestingly, a SNP in very high (LD) with this SNP (rs11685299; LD R^2^ = 0.99) is genome-wide significant for association with SCZ (29). The BP allele of a SNP in high LD with both of these SNPs (rs11681451) is also associated with decreased expression of CUL3, this time in three brain regions (Cortex P = 1.81×10^−6^, Hippocampus P = 1.95×10^−5^, Caudate P = 2.33×10^−5^; Supplementary Table 5).

The gene *PSMD2* encodes a subunit of the 26S proteasome, which degrades NRF2 (28). At *PSMD2*, the BP allele of rs843355 is associated with decreased expression of *PSMD2* in various immune cells in the blood, including Blood-Neutrophils CD16+ (P = 1.38×10^−6^) and Blood-Monocytes (P = 1.79×10^−6^) (Supplementary Table 5).

Similarly to *PSMD2,* the *PSMB10* gene also encodes a proteasomal subunit involved in NRF2 degradation (28). At *PSMB10*, the BP allele of rs5923 is associated with both increased and decreased expression of *PSMB10* in various blood immune cells (Increased: Blood P = 4.2×10^−9^, Blood-Monocytes CD14+ P = 5.11×10^−9^, Blood-Neutrophils CD16+ P = 2.94×10^−7^; Decreased: Blood-Monocytes CD14+ P = 9.56×10^−7^, Blood-Neutrophils CD16+ P = 8.21×10^−6^).

For the *mRNA Splicing - Minor Pathway*, six genes satisfied the criteria of having gene P values < 1×10^−2^ for BP and < 1×10^−3^ for chronotype: *splicing factor 3b subunit 1* (*SF3B1*)*, splicing factor 3b subunit 1* (*SF3B2*)*, RNA polymerase II, I And III subunit H* (*POLR2H*)*, small nuclear ribonucleoprotein U5 Subunit 200* (*SNRNP200*) and *pre-mRNA processing factor 6* (*PRPF6*).

*SF3B1* encodes subunit 1 of the splicing factor 3b protein complex (SF3b) which is a component of the U12 small nuclear ribonucleoprotein (snRNP) of the minor spliceosome and a component of the U2 snRNP of the major spliceosome. At *SF3B1*, the BP allele of the SNP most associated with BP (rs55775495) is associated with increased expression of SF3B in the central nervous system (P = 2.68×10-15), peripheral nervous system (P = 1.13×10^−6^), brain cerebellum (P = 1.30×10^−6^) and brain caudate (P = 2.19×10^−6^). Interestingly, the SNP rs6434928, which is in very high LD (R^2^ = 0.88) with this SNP is genome-wide significant for association with SCZ (30) and has almost the exact same eQTL results. *SF3B2* encodes subunit 2 of SF3b. The BP allele of rs2270448 is associated with decreased expression of *SF3B2* in the brain cerebellum (P = 1.28×10^−5^).

*POLR2H* encodes a subunit shared by the three eukaryotic polymerases. The BP allele of rs62287507 is associated with increased expression of POLR2H in blood (P = 2.33×10^−6^). *SNRNP200* encodes an RNA helicase that comprises one of the U5 snRNP-specific proteins. At *SNRNP200*, the BP allele is associated with increased expression in blood neutrophils CD16+ (P = 2.02×10^−11^). *PRPF6* encodes a protein that acts as a bridging factor between U5 and U4/U6 snRNPs in formation of the spliceosome. The encoded protein can also bind androgen receptor, providing a link between transcriptional activation and splicing. The BP allele of the most significant SNP at this loci, rs3829704, is associated with decreased expression of PRPF6 in blood cells (Lymphocyte P = 1.29×10^−31^, Blood P = 3.95×10^−11^, Blood-T cell CD4+ activated P = 1.48×10^−7^) and decreased expression of PRPF6 in the brain (Cortex P = 1.2×10^−6^; Caudate, P = 6.0×10^−6^).

#### Differential Expression Analysis

We next investigated if genes within the *mRNA Splicing – Minor Pathway* and *KEAP1 NRF2* pathway are differentially expressed between BP cases and controls. We used PsychEncode differential expression data based on postmortem dorsolateral prefrontal cortex samples from 144 BP cases and 899 healthy controls. Two genes within the mRNA Splicing – Minor Pathway were found to be differentially expressed. These are *Serine and Arginine Rich Splicing Factor 6* (*SRSF6*; log2FC = −0.07; P = 2.0×10^−4^; FDR = 0.01) and *Small Nuclear Ribonucleoprotein Polypeptides B and B1* (*SNRPB*; log2FC = 0.08; P 4.4×10^−4^= FDR = 0.02). Four differentially expressed genes were observed in the *KEAP1 NRF2* pathway. These are *Sestrin 1* (*SESN1*; log2FC = 0.07; P = 2.8×10^−4^; FDR = 0.02), *cyclin dependent kinase inhibitor 1A* (*CDKN1A* log2FC = 0.32; P = 2.51×10^−4^; FDR = 0.02), *AKT serine/threonine kinase 1* (*AKT1*; log2FC = 0.04; P = 6.8×10^−4^; FDR = 0.03) and *MAF BZIP Transcription Factor G* (*MAFG*; log2FC = −0.05; P = 4.9×10^−4^; FDR = 0.02). Interestingly, three of these genes are present in *Regulation of NRF2 by KEAP1*, which had a higher average R^2^ per SNP than the *KEAP1 NRF2* pathway itself (Figure 4b). However, none of these gene-sets had a statistically significant enrichment of differentially expressed genes (*Regulation of NRF2 by KEAP1* P= 0.09; *KEAP1 NRF2* P = 0.5; *mRNA Splicing – Minor Pathway* P =0.5).

## Discussion

Using genome-wide polygenic score analysis, we have found that polygenic scores for AUT, SCZ and BP could predict an evening chronotype and polygenic scores for all four NDPC phenotypes could predict insomnia status in UKB. Pathway-based polygenic score analysis identified the *NRF2 KEAP1* and *mRNA splicing minor* pathways as being enriched for genetic variation overlapping between chronotype and BP. For the *NRF2 KEAP1* pathway, the signal is enriched in the subset of genes that function with KEAP1 to regulate *NRF2* expression. Examination of eQTL data pointed to BP associated SNPs being associated with expression changes of many genes within the highlighted pathways to which they map. Interestingly, three of the ten eQTL SNPs are genome-wide significant for SCZ (Supplementary Table 5).

The genome-wide polygenic score results we report support previously published genetic correlation results. AUT and SCZ have previously been reported to exhibit a negative genetic correlation with a morning chronotype, as demonstrated by linkage disequilibrium score regression (31,32). Using a different method and data, our study provides further evidence for these findings, as well as demonstrating a positive association between the BP PGS and an evening chronotype. Additionally, our results confirm that insomnia is genetically correlated with ADHD, SCZ and BP (33–35), the strongest relationship being between ADHD and insomnia. We also report a negative genetic correlation between AUT and insomnia, whereas, in previous studies this relationship was not statistically significant (31).

BP polygenic scores created using SNPs that map to the *KEAP1 NRF2* and *mRNA splicing-minor* Reactome pathways were associated with chronotype, over and above the background signal. The majority of the variance explained by the *KEAP1 NRF2* gene-set is concentrated within genes belonging to the subpathway, *Regulation of NRF2 by KEAP1*. It must be noted that the R^2^ values of these pathway-based PGSs are low. This is partly to be expected, given the small numbers of genes in these pathways. Clear enrichment of explained variance by the *mRNA splicing – minor* and *KEAP1 NRF2* pathways are observed when plotting the average variance explained per SNP for these pathways in comparison to the average variance explained per SNP by PGSs created using all genes based on BP genetic variation (Figure 4b).

A sub-pathway of *KEAP1-NRF2*, *Nuclear Events Mediated by NRF2*, ranked very highly in predicting insomnia status, based on BP and SCZ genetic variation. This did not reach statistical significance, possibly due there being less insomnia samples (N=239,918) than chronotype samples (N=409,630), and therefore reduced power. NRF2 is a transcription factor that, under normal conditions, is bound by KEAP1 in the cytoplasm as part of the CUL3-KEAP1 E3 ubiquitin ligase complex, which facilitates the ubiquitination of NRF2, marking it for proteasomal degradation. When cells are exposed to oxidative stress, toxins, bacteria, viruses or other forms of cellular damage, KEAP1 cysteine residues are modified, disrupting its binding affinity for NRF2. As a result, NRF2 stabilises and translocates to the nucleus where it binds antioxidant response elements (AREs) in the promoter regions of genes encoding antioxidant enzymes and detoxification proteins (36). Of particular interest regarding sleep and circadian rhythms, NRF2 has also been observed to undergo rhythmic regulation by the two circadian clock genes, *BMAL1* and *CLOCK*, in myeloid cells. These clock genes promote *NRF2* expression by binding to the NRF2 E-box domain (37). Through these pathways, NRF2 mediates coupling between oscillations in redox balance and circadian timekeeping mechanisms, with this loop serving to optimise the timing of circadian and seasonal timing of antioxidant responses (38,39).

Elevated oxidative stress has been consistently documented in numerous NDPCs (including BP), and this contributes to the concurrent neuroinflammation observed in these conditions (40,41). The increased expression of NRF2 target genes help cells mitigate oxidative stress, improve mitochondrial function and reduce inflammation, all of which are involved in the pathology of NDPCs (36,42). NRF2 also acts as a negative regulator of the NFκB signalling pathway, while the NFκB pathway reciprocally negatively regulates NRF2 signalling. This interaction helps maintain control over basal inflammation and oxidative stress levels (43). When the NRF2 pathway becomes saturated or is deficient, activation of the NFkB pathway is heightened, which leads to an increased production of pro-inflammatory cytokines (36). Given this relationship, it is therefore interesting that the *KEAP1 NRF2* and *Nuclear Events Mediated by NRF2* pathways clustered together with the *FCERI mediated NF-kB activation* pathway competitive P value heatmap of the top four pathways across all phenotype pairs (Figure 3).

Studies in a seasonal animal model (Meduka fish) that exhibits a constitutive depressive winter phenotype have shown that NRF2 signalling pathways were inactivated in winter photoperiods, and that this was associated with a depressive phenotype that could be reversed by treatment with NRF2 activators (44). These functions of NRF2 signalling in regulating seasonal changes in behaviour are significant since seasonality is such an important clinical feature of human NDPCs. For example, the prevalence of major depressive disorder and seasonal affective disorder show seasonal variation and the oscillations in mood and metabolic function that characterise BP recur with a seasonal rhythm (45). Furthermore, people with BP have higher self-reported seasonality scores (46) and altered sensitivity to changes in day light (47). Chronotype also links seasonality and BP, as evening chronotype is associated with increased self-reported seasonality (48) and with the presence and severity of signs of BD (49).

Given the beneficial antioxidant and anti-inflammatory roles of NRF2, it may be hypothesized that NRF2 is downregulated in NDPCs. This was observed in animal models of depression, where lower levels of NRF2 and KEAP1 were reported in the prefrontal cortex and the dentate gyrus and Cornu Ammonis 3 (CA3) regions of the hippocampus, along with increased blood levels of inflammatory cytokines (50). Similar results were seen using human postmortem samples, where protein levels of KEAP1 and NRF2 in the parietal cortex of major depressive disorder, schizophrenia, and bipolar disorder samples were significantly lower than controls. Indeed, there is evidence supporting the potential of natural and synthetic molecules, including melatonin, that elevate NRF2 levels to be used as treatments for psychiatric conditions (36,42).

However, there is some disagreement in the literature, with some studies reporting that NRF2 signalling is upregulated in NDPCs. For example, Lukic et al. (2014) reported increased abundance of NRF2, KEAP1, and NF-κB within the cytoplasm of peripheral blood mononuclear cells of MDD patients (51). This lack of consistent direction, in terms of up-or down-regulation of NRF signalling, is reflected in our eQTL results, where BP genetic variation is associated with inconsistent changes in expression of genes within the NRF2 KEAP gene-set. For example, the CUL3 gene encodes the cullin 3 protein, which is a core component of ubiquitin ligase complex which ubiquitinates NRF2 and the PSMD2 gene encodes a subunit of a proteasome that degrades NRF2. Therefore, CUL3 and PSMD2 function together to degrade NRF2, yet BP eQTLs within these genes are associated with expression changes that are in opposite directions. Additionally, the four genes within the NRF2 KEAP1 gene-set that are differentially expressed between BP cases and controls, based on PsychEncode differential expression data, are upregulated.

The other pathway highlighted by our study is the *mRNA-splicing minor pathway*. Splicing of mRNA refers to the removal of introns from pre-mRNA transcripts, a process performed by the spliceosome. The spliceosome comprises distinct complexes responsible for various mRNA splicing steps, composed of both proteins and small nuclear RNAs. While the majority of splicing events are carried out by the major spliceosome, a subset of introns, known as minor introns, necessitates the involvement of the minor spliceosome for their excision (52). Minor splicing is less efficient than major splicing, this reduced efficiency has been proposed to play a role in the regulation of genes containing minor introns, as minor intron containing transcripts are retained in the nucleus until the minor spliceosome components become available. Of relevance to NDPCs, the cerebral cortex is one of the tissues with the highest expression of genes containing minor introns (53).

The importance of minor intron splicing is evident by the fact that minor-intron containing genes are highly conserved and thirteen diseases have been linked to mutations in minor splicing components, including the neurodegenerative diseases, amyotrophic lateral sclerosis, and spinal muscular atrophy (52,54,55). A study by Buel and colleagues found oscillations in many components of the major spliceosome pathway mirror that of the *clock and bmal1* transcripts (56), indicating that the major spliceosomal pathway is at least partly regulated by the circadian clock. This is interesting in relation to the minor spliceosome too, given the overlap in these pathways. Interestingly, the biologically related pathway, *Processing of Capped Intron-Containing Pre-mRNA*, ranked as the second highest performing pathway PGS in the prediction of insomnia based on ADHD genetic variation. This pathway clustered together with *mRNA-splicing minor pathway* in the heatmap in Figure 3.

Neither the *NRF2 KEAP1* or the *mRNA-splicing minor* pathways were found to be enriched in genes differentially expressed between BP cases and controls in the *post-mortem* dorsolateral prefrontal cortex. It could be that these genes are differentially expressed in a different tissue type to the one we investigated or that the shared genetic variation we have identified only applies to a subset of individuals, and differential expression of these genes would only occur for this subset. A future analysis could select BP cases with high *NRF2 KEAP1 / mRNA-splicing minor* PGSs and restrict the differential analysis to just these cases (this would require genotype and RNA-sequencing data on BP cases and controls).

We implemented MAGMA gene-set analysis to investigate if the *NRF2 KEAP1* and *mRNA splicing minor* pathways could be identified using standard single phenotype pathway enrichment methods. Neither the *NRF2 KEAP1* and *mRNA splicing minor pathways* were enriched for genetic variation individually associated with BP, chronotype or SCZ (Supplementary Table 6). This indicates that these pathways are enriched for the genetic variation shared between these two phenotypes and are not enriched for the genetic variation associated with each individual phenotype.

There are some limitations to the pathway-based PGS method we have implemented. Firstly, we mapped SNPs to genes solely based on the physical distance of SNPs to genes. While, it has been reported that the majority (76%) of GWAS SNPs affect the closest gene (57), SNPs can be located in regulatory elements up to hundreds of megabases away from the causal gene. Such distal causal genes were not included in our analysis. Another limitation is that polygenic score analysis cannot distinguish between horizontal versus vertical pleiotropy i.e. whether genetic variation within the *NRF2 KEAP1* and *mRNA splicing minor* pathways independently contribute to both BP and an evening chronotype, or whether genetic variation within these pathways contribute to an evening chronotype, which in turn, causes BP, or vice versa.

This is the first study to investigate the biological pathways enriched in genetic variation overlapping between NDPC and sleep phenotypes, using pathway-based PGS analysis in a global, rather than hypothesis-based design. We used the largest available datasets from GWASs of ADHD, AUT, BP as our discovery datasets, and our target dataset consisted of up to 409,630 samples from UKB.

Our findings of shared genetic variation between chronotype and BP in a pathway that is known to regulate circadian rhythms and seasonality in animals (*NRF2 KEAP1*) in addition to the strongly seasonal nature of the clinical features and prevalence of BP, are further evidence to support the hypothesis that disrupted circadian rhythm and seasonal adaptive responses underlie the pathology of BP (58). Furthermore, our findings highlight the *mRNA splicing-minor* pathway, which has also been shown to be at least partly regulated by the circadian clock. Thus, this study provides further evidence to support the circadian clock hypothesis of BP disorder.

## Methods

### Target Data for Polygenic Score Analysis – UK Biobank

#### Quality Control (QC)

Genotype data was collected, processed and imputed by UKB (59). We accessed and analysed the data through the UKB Research Analysis Platform (Application ID: 71469). UKB research participants were excluded based on the following criteria: not being of Caucasian ancestry (data-field: 22006), containing a sex chromosomal aneuploidy (data-field: 22019), having a high SNP missingness and/or having unusually high or low heterozygosity (data-field: 22027), having ten or more third-degree relatives (data-field: 22021) and performing shift work (data-field: 826). Further samples were removed based on relatedness using the UKB supplied relatedness file, which lists pairs of individuals related up to a third degree. One individual from each pair was removed without removing samples from pairs where a sample had already been removed. A total of 487,409 samples passed these quality control measures. To minimize any potential overlap between UKB and PGC SCZ and BP cohorts, participants who had received SCZ or BP diagnoses were excluded from the construction of the SCZ PGS or BP PGS, respectively. Such samples were identified through self-reported diagnoses (data-field: 20544) and linked health records reporting international classification of disease (ICD10) codes, F200-F209 for SCZ and F310-F319 for BP (data-field: 41270). This resulted in a further removal of 1,223 SCZ samples and 1,866 BP samples, in the construction of the SCZ and BP PGSs, respectively.

SNPs were excluded based on the following criteria: imputation quality score (INFO) < 0.7, proportion of missing genotypes > 0.02, minor allele frequency (MAF) < 0.005, Hardy–Weinberg equilibrium (HWE) < 1×10^−6^. Duplicate SNPs were removed using Plink2 with the flag “--rm-dup force-first”. Plink2 binary file-sets were created using the “--make-bed” flag and chromosome specific files were merged using the Plink2 “--merge-list” flag. Finally, ambiguous SNPs and SNPs not present in the corresponding neuropsychiatric GWAS were removed.

#### Phenotype Generation

Two UKB phenotypes were used for this study: chronotype (“Morning/evening person (chronotype)”; data-field 1180) and insomnia (“sleeplessness/insomnia”; data-field 1200). These data points were gathered as part of a questionnaire participants completed on touchscreen computers at UKB assessment centres.

For chronotype, participants were asked the question, “do you consider yourself to be”?, and they had six optional answers, which we encoded as a quantitative eveningness phenotype: “Definitely an ‘evening’ person” was encoded as 4, “More an ‘evening’ than a ‘morning’ person” was encoded as 3, “More a ‘morning’ than an ‘evening’ person” was encoded as 2, “Definitely a ‘morning’ person” was encoded as 1, “Prefer not to answer” and “Do not know” were both encoded as missing. This resulted in 409,630 post-QC samples with chronotype data.

For insomnia, participants were asked the question, “do you consider yourself to be”? and had four optional answers, which we encoded in a binary fashion: “never/rarely” was encoded as 0, “usually” was encoded as 1 and “sometimes” and “Prefer not to answer” were both encoded as missing. This resulted in post-QC 239,918 samples with insomnia data.

#### Correcting for Confounders

The UKB phenotypes were corrected for the effects of confounders by taking the residuals of regression models with the UKB phenotype as the dependent variable and the confounders as the independent variables. For the binary insomnia phenotype, a logistic regression model was used, while a linear regression model was used for the quantitative chronotype phenotype. The residuals of these models, representing the variation in phenotype left unexplained by the effects of confounders, were assigned as the corrected phenotypes. The fourteen confounders corrected for were: age, sex, UKB assessment centre, genotyping batch and genetic principal components (PCs) 1-10.

### Discovery Data for Polygenic Score Analysis – GWAS Summary Statistics

Discovery data for polygenic score analysis included summary statistics from the latest and largest published GWAS of AUT (18,381 cases and 27,969 controls from a Danish population (31)), ADHD (38,691 cases and 186,843 controls from European, Danish and Icelandic populations (33)), SCZ (74,776 cases and 101,023 controls from European, East Asian, African American and Latino populations (30)) and BP (41,917 cases and 31,358 controls from a European population (35)).

### Genome-Wide Polygenic Score Analysis

Effect sizes for all imputed SNPs were estimated using SBayesRC based on GWAS summary statistics. SBayesRC models SNP effects using a mixture of normal distributions that differ in their variances and uses SNP annotation information to select which of these distributions best model SNP effect sizes. This allows for SNP effect probability distributions to differ across annotation groups and has been shown to perform better than other popular PGS methods (24). Inputs to SBayesRC include GWAS summary statistics, LD European reference data and data containing 96 functional annotations for 8 million imputed SNPs (the latter two data files were downloaded from github.com/zhilizheng/SBayesRC).

The first step involves converting GWAS summary statistics to COJO format, and converting odds ratios to beta values using log(OR) when beta values were not provided. Subsequently, the SBayesRC tidy function was run to filter SNPs based on inconsistent alleles between the GWAS summary statistic and LD data and per SNP sample sizes less than three standard deviations of the mean. The SBayesRC impute function was implemented to impute the GWAS summary data based on LD. This resulted in 7,356,519 SNPs for each phenotype. The main SBayesRC function was then run to estimate SNP effects. These SNP effects were provided to the--score command in Plink1.9 to calculate PGSs for all individuals in the UKB test dataset (60). Performance metrics were calculated in R by running linear regressions with UKB chronotype or insomnia status as the dependent variable and the SCZ, BP, AUT or ADHD PGS as the independent variables. P values and variance explained (R^2^) were obtained using R’s summary function. Confidence intervals were created using the ci_rsquared function as part of the confintr R package (https://github.com/mayer79/confintr).

### Pathway-Based Polygenic Score Analysis

#### Pathway data

Pathway data for pathway based PGS analysis was sourced from Reactome, an open-source, manually curated and peer-reviewed pathway database (61). We used all *Homo sapien* pathways that contained greater than 50 genes, to remove overly specific pathways, and less than 500 genes, to remove overly broad pathways and limit the computational intensity of pathway-based PGS analysis. This resulted in 451 pathways to test.

#### Pathway-based polygenic score analysis using PRSet

Polygenic score analysis was performed using the tool, PRSet (62). Inputs to PRSet included Reactome pathway data, published GWAS summary statistics to be used as discovery data, UKB files in PLINK binary format (.bed, .bim and .fam) to be used as target data and a GTF file for the genome build, GRCh37. SNPs with a reported P > 0.5 from the corresponding GWAS were removed. This resulted in final SNP numbers of N = 2,374,077 for the AUT PGS, N = 2,187,591 for the ADHD PGS, N = 2,525,546 for the SCZ PGS and N = 2,416,496 for the BP PGS.

Pathway-based PGS analysis in PRSet involves three steps. The first step is to map SNPs to genes by genomic position using GRCh37 start-stop coordinates extended 35kb upstream and 10kb downstream to include potential regulatory elements. The second step is to perform pathway specific LD clumping (parameters used:--clump-kb 250kb--clump-p 1--clump-r2 0.1). Clumping is performed for each pathway independently to retain the genetic signal for each pathway i.e to prevent a SNP outside of the pathway being assigned as the lead clumped SNP. The third step involves calculating pathway specific PGSs for each individual, by summing up, for each SNP, the number of minor alleles at that SNP multiplied by the GWAS effect size. Finally, performance metrics (R^2^ and P value) are calculated for each pathway. Competitive analysis was performed by creating 5,000 background polygenic scores, each constructed by randomly sampling the same number of post LD clumped SNPs as contained within gene-set being tested and conservatively including the gene-set being tested as one background set. A competitive P value is created using a formula that counts the number of times the P value for a background gene-set is less than the P value for the gene-set being tested and dividing by the number of permutations (62). A disadvantage of this method is that the lowest possible P value is 1 / (number of permutations + 1). Therefore, gene-sets that reached the lowest possible competitive P value at 5,000 permutations were re-analysed with a higher number of permutations. Confidence intervals were calculated for the top performing pathways in R using the same method as for the genome-wide PGSs.

Multiple testing correction was performed for pathway-based PGS analysis by calculating a Bonferroni corrected P value threshold, where 0.05 was divided by the number of tests performed (four GWAS phenotypes x 451 pathways x 2 UKB phenotypes = 3,608).

### Post-hoc analysis

#### MAGMA gene-set analysis

Gene-sets that were statistically significant in the PGS analysis, and the genes within them, were tested for enrichment of common genetic variation using region-based multi-marker analysis of genomic annotation (MAGMA; http://ctg.cncr.nl/software/magma; (63) and summary statistics from published GWAS on chronotype (69,369 cases and 236,642 controls of British ancestry; 3) and BP. An analysis involves three steps. First, in the annotation step, SNPs with available GWAS results are mapped on to genes (GRCh37/hg19 start-stop coordinates +35kb/-10kb). Second, in the gene analysis step, gene P values are computed for each GWAS dataset. This gene analysis is based on a multiple linear principal components regression model that accounts for LD between SNPs in each gene, number of SNPs in each gene, inverse of the mean minor allele count of variants in each gene and the GWAS sample size. The European panel of the 1000 Genomes data was used as a reference panel for LD. Third, a competitive GSA based on the gene P values, also using a regression structure, was used to test if the genes in each gene-set were more strongly associated with either phenotype than other genes in the genome.

#### eQTL Analysis

Expression quantitative trait loci (eQTL) data was examined using QTLbase2 (27), a database that aggregates published genome-wide QTL summary statistics across more than 95 tissue and cell types. We reviewed eQTL data related to genes found within the gene-sets prioritised by pathway-based PGS analysis. Specifically, we considered genes that achieved significance levels of P < 1×10^−2^ for BP and P < 1×10^−3^ for chronotype in the gene-based MAGMA analysis. This was to identify genes individually enriched for BP and chronotype genetic variation, as these are the genes most likely contributing to the significant pathway-based PGS results and therefore of interest. Since these genes were already highlighted as being of interest through pathway-based PGS analysis, it was not necessary that they were individually statistically significant in the MAGMA analysis.

The most statistically significant SNP at each gene locus was searched in QTLbase to identify if it had been associated with expression changes of the mapped gene in biologically relevant tissues or cell types, including those related to the brain, central nervous system, peripheral nervous system, blood, or immune systems. If the top SNP at a locus did not show any eQTL associations, we investigated other SNPs with similarly low P values at that locus. It was assumed that the SNPs demonstrating the highest statistical significance for association with BP would also contribute to chronotype prediction, given that BP effect sizes were used to compute the pathway-based PGSs.

#### Differential Expression Analysis

Differential expression summary data was downloaded as Supplementary Table 1 from Gandal et al. (2018). This is a PsychENCODE Consortium study that performed RNA-sequencing, followed by differential expression analysis, of samples isolated form the postmortem dorsolateral prefrontal cortex samples for 144 BP cases and 899 controls. Specific details of the analysis performed are provided in the paper (64)

More specific methodology details and code for all analyses are provided here: https://github.com/laurafahey02/ Polygenic_Score_Analyses.

## Data Availability

UK Biobank genotype and questionnaire data can be accesssed via the UK Biobank research analysis platform (RAP): https://ukbiobank.dnanexus.com/landing. The Research Analysis Platform is open to researchers who are listed as collaborators on UKB-approved access applications. All results produced in the study are available in the manuscript or supplementary tables.

## Acknowledgements

This project has received funding from the European Research Council (ERC) under the European Union’s Horizon 2020 research and innovation programme (grant agreement No 950010). We would like to thank Dr Cathy Wyse for discussions related to chronobiology and for reviewing a draft of the manuscript.

